# Causal association between serum bilirubin and ischemic stroke: Multivariable Mendelian Randomization

**DOI:** 10.1101/2023.12.20.23300340

**Authors:** Jong Won Shin, Keum Ji Jung, Heejin Kimm, Sun Ha Jee

**Author notes:** Both authors contributed equally as first authors (Jong Won Shin, Keum Ji Jung). **Correspondence:** Sun Ha Jee, PhD, MPH Department of Epidemiology and Health Promotion, Graduate School of Public Health, 50-1 Yonsei-ro, Seodaemun-gu, Seoul 03772, Korea.

## Abstract

**BACKGROUND:** Past studies have mainly focused on total bilirubin levels, and have not clearly distinguished between direct and indirect bilirubin, a subgroup of bilirubin. In this study, the differences between these subgroups were clearly examined, and the causal association with ischemic stroke was examined in more detail.

**METHODS:** Utilizing Two sample Multivariable Mendelian randomization (MVMR) analyses, summary data for bilirubin were extracted from the KCPS-II (Korean Cancer Prevention Study-II; n=159,844) and the KoGES (Korean Genome and Epidemiology Study; n=72,299), while ischemic stroke data were derived from the BBJ (Bio Bank Japan; n=201,800).

**RESULTS:** The crude two-sample MR analysis revealed a significant negative association between total bilirubin and ischemic stroke in KoGES data (odds ratio [OR], 0.86; 95% confidence interval [CI], 0.75-0.98). Subsequent bivariable MR analyses, controlling for lipid profile, also showed significant results. In KCPS-II data, direct bilirubin showed significance in both crude (0.65, 0.43-0.97) and bivariable analyses, while indirect bilirubin demonstrated significant associations in MVMR analyses (0.76, 0.59-0.98), emphasizing its role in mitigating the risk of ischemic stroke.

**CONCLUSIONS:** Our study establishes a causal association between genetically determined levels of serum bilirubin (total, direct, and indirect) and a reduced risk of ischemic stroke in an Asian population. Notably, the protective effect was predominantly associated with indirect bilirubin. The findings highlight the significance of considering bilirubin subgroup in understanding the mechanisms underlying endogenous antioxidants and its impact on ischemic stroke.

## INTRODUCTION

Stroke is a form of oxidative stress-related disease.^1,2,3^ Serum bilirubin levels are widely reported to be inversely related to stroke risk in both Western populations^4,5^ and Asian populations.^6,7^ These studies have been based on serum total bilirubin levels. In other words, it is a result that does not take into account the specific levels of direct bilirubin (conjugated bilirubin) and indirect bilirubin (unconjugated bilirubin) that make up the total bilirubin. In fact, indirect bilirubin levels account for a large proportion of the subgroups of total bilirubin (reference values - total bilirubin: 0.3-1.0 mg/dl, direct bilirubin: 0-0.3 mg/dl, indirect bilirubin: 0.2-0.8 mg/dl). Aged red blood cells are broken down into heme and globin, and then heme is broken down into iron (Fe2+) and biliverdin. In the process of oxidation to biliverdin (Figure 1), the level of indirect bilirubin acts as a powerful endogenous antioxidant.^8,9^

**Figure 1.**
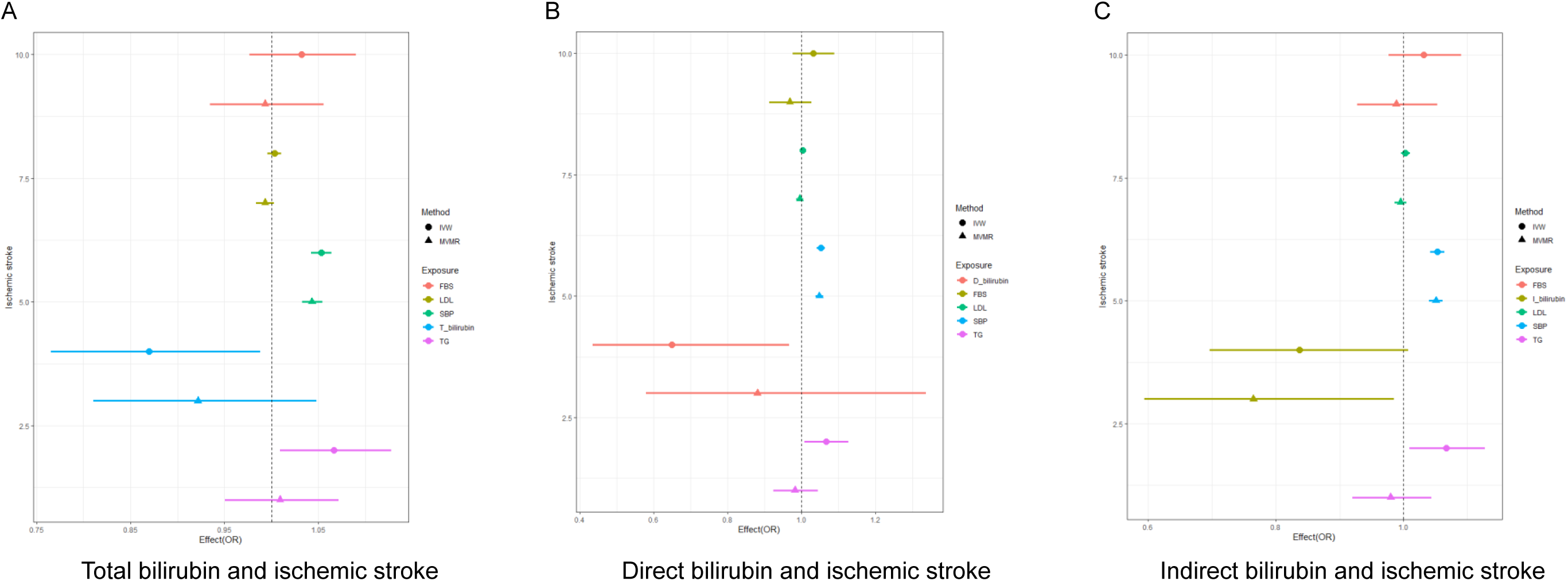
Causal association between total, direct, and indirect bilirubin and ischemic stroke using MVMR

The First study on total bilirubin and stroke occurrence using genetic information was a case-cohort design of 806 stroke patients that occurred in a subcohort of 4,793 subjects in the KCPS-II (Korean Cancer Prevention Study-II). This study, a one-sample mendelian randomization (MR) analysis, showed a negative association between total bilirubin and total stroke (HR=0.63, 95% CI 0.30–1.36), but did not show statistically significant results (p=0.240).^10^ The second study performed a two-sample MR analysis of total bilirubin based on KoGES (Korean Genome and Epidemiology Study; n=25,406) and KCPS-II (n=14,541) biobank data.^10^ The results provided evidence for a significant causal association between total bilirubin levels and reduced risk of stroke in the Korean population. In particular, the association was greater for ischemic stroke (OR=0.302) than for total stroke (OR=0.481).^11^ However, considering only serum total bilirubin, there was insufficient specific evidence to suggest a reduction in stroke risk through bilirubin’s role as an endogenous antioxidant.

## OBJECTIVES

1. We divided bilirubin into subgroups to determine whether each subgroup plays a role as an endogenous antioxidant.
2. We also investigated whether it was possible to suggest the significance of causal associations when adjusting for several genetic variables, which was not possible in existing MR studies.

## METHODS

We analyzed total, direct, and indirect bilirubin data from KoGES (n=72,299)^12^ and KCPS-II (n=159,844) biobanks^13^ and ischemic stroke from BBJ (BioBank Japn; n=201,800).^14^ data was used to perform Two Sample Multivariable MR (MVMR). In other words, MVMR was performed to control blood pressure, fasting blood sugar, and serum lipid levels related to stroke.

### Genetic instruments for serum bilirubin (G-X)

A genetic instrumental variable for serum bilirubin was identified using two Korean biobanks (KCPS-II and KoGES).^12,13^ The selection of instrumental variables for MR analysis was based on the following criteria: First, cases with a *p-value* smaller than the genome-wide significance level identified in the study (*p-value* <5×10^-8^). The second, minor allele frequency (MAF) is greater than 0.01. The third, single nucleotide polymorphism (SNP)s with linkage disequilibrium (LD) relationship were excluded (clumping criteria: r^2^ 0.001 standard). Finally, palindromic (SNP) was excluded from the analysis (MAF > 0.42).

### Genetic associations of SNPs with ischemic stroke (G-Y)

Summary data used for ischemic stroke were obtained from BBJ.^14^ BBJ is a biobank composed of 201,800 patient data centered on 66 hospitals nationwide between 2003 and 2008.

### Mendelian randomization

In this study, G-X data on exposure as two-sample MR came from KCPS-II and KoGES. And G-Y data for outcome came from BBJ. The coefficient was estimated using the inverse variance weighted (IVW) method under the assumption that all selected SNPs were valid. At this study, coefficient was calculated by calculating the coefficient of each SNP according to the wald ratio method and combining it using the IVW method.

Multivariable MR (MVMR) analysis was performed controlling for genetic variables of triglyceride (TG), cholesterol (LDL; low density lipoprotein, HDL; high density lipoprotein), systolic blood pressure (SBP), and fasting blood sugar (FBS), which are noted as major causes of stroke. To ensure the validity of instrumental variables, the F-value of each variable was presented in each model.

Analyses were conducted using the Two Sample MR package in R, version 3.6.0 (R Project for Statistical Computing).

## RESULTS

This study divided bilirubin into total, direct, and indirect bilirubin and analyzed Mendelian randomization for each of them for their association with the risk of ischemic stroke. In this study, summary data for bilirubin was extracted from KoGES and KCPS-II, respectively, and summary data for ischemic stroke was used from BBJ data.

Table 1 shows crude two-sample MR and MVMR results on the effect of total bilirubin on ischemic stroke. It showed a significant negative association that crude two sample MR analysis between total bilirubin and ischemic stroke in KoGES data (OR, 0.86, 95%CI, 0.75-0.98). In addition, MVMR, controlling for LDL, HDL, and TG, respectively, showed significant results. Additionally, there are borderline significant results in the crude two-sample MR results and in the model controlling LDL from KCPS-II. However, MVMR, controlling for HDL and TG, showed significant results. In the remaining models, a negative association between total bilirubin and ischemic stroke was observed, but it was not significant (Figure 1-A). The F-statistic of total bilirubin and other variables used in the model in Table 1 were all above 10 except for systolic blood pressure (SBP), and the assumption as an instrumental variable was satisfied (Supplementary Table 1).

**Table 1.** Causal effect of total bilirubin on ischemic stroke.

|  | KoGES (total bilirubin), BBJ (ischemic stroke) |  | KCPS-II (total bilirubin), BBJ (ischemic stroke) |  |
| --- | --- | --- | --- | --- |
|  | IVW method, OR (95% CI) | P | IVW method, OR (95% CI) | P |
| Crude two sample MR | 0.86 (0.75-0.98) | 0.0294 | 0.87 (0.77-0.99) | 0.0652 |
| MVMR |  |  |  |  |
| Adjusted for LDL | 0.86 (0.76-0.96) | 0.0097 | 0.89 (0.79-1.01) | 0.0669 |
| Adjusted for HDL | 0.82 (0.73-0.93) | 0.0020 | 0.88 (0.79-0.99) | 0.0326 |
| Adjusted for TG* | 0.99 (0.99-1.00) | 0.0545 | 0.87 (0.78-0.98) | 0.0174 |
| Adjusted for LDL and HDL | 0.96 (0.75-1.23) | 0.7520 | 0.95 (0.81-1.12) | 0.5259 |
| Adjusted for LDL and TG | 0.97 (0.76-1.23) | 0.7910 | 0.91 (0.79-1.05) | 0.2009 |
| Adjusted for HDL and TG | 0.84 (0.65-1.08) | 0.1751 | 0.94 (0.82-1.09) | 0.4217 |
| Adjusted for LDL, HDL and TG | 0.99 (0.78-1.25) | 0.9070 | 0.94 (0.82-1.08) | 0.3682 |
| Adjusted for LDL, HDL, TG and SBP | 0.93 (0.76-1.15) | 0.5138 | 0.95 (0.84-1.08) | 0.4658 |
| Adjusted for LDL, TG and SBP | 0.87 (0.64-1.19) | 0.3891 | 0.92 (0.81-1.05) | 0.2435 |
| Adjusted for LDL, TG, SBP and FSG | 0.90 (0.68-1.20) | 0.4877 | 0.92 (0.81-1.05) | 0.2136 |
| Adjusted for LDL, TG, and FSG | 0.83 (0.65-1.05) | 0.1249 | 0.91 (0.79-1.04) | 0.1720 |

KoGES, Korean Genome Epidemiologic Study; KCPS-II, Korean Cancer Prevention Study-II; BBJ, Biobank of Japan; IVW, inverse variance weighted method; OR, odds ratio; CI, confidence interval; MVMR, multivariable mendelian randomization; LDL, low density lipoprotein; HDL, high density lipoprotein; TG, triglyceride; SBP, systolic blood pressure; FBS, fasting serum glucose

Table 2 shows the crude two sample MR and MVMR results on the effect of direct bilirubin on ischemic stroke. KoGES data analysis showed a significant negative association only in crude two-sample MR (p=0.0493). However, KCPS-II data analysis showed significant results in crude two sample MR, and also showed significant results in MVMR with LDL, HDL, and TG controlled respectively. The remaining model showed a negative relationship, but it was not significant (Figure 1-B). The F values of direct bilirubin and control variables used in the model in Table 2 were all above 10 except for SBP, and the assumption as an instrumental variable was satisfied (Supplementary Table 2).

**Table 2.** Causal effect of direct bilirubin on ischemic stroke.

|  | KoGES (direct bilirubin), BBJ (ischemic stroke) |  | KCPS-II (direct bilirubin), BBJ (ischemic stroke) |  |
| --- | --- | --- | --- | --- |
|  | IVW method, OR (95% CI) | P | IVW method, OR (95% CI) | P |
| Crude two sample MR | 0.67 (0.45-1.00) | 0.0493 | 0.65 (0.43-0.97) | 0.0335 |
| MVMR |  |  |  |  |
| Adjusted for LDL | 0.94 (0.55-1.60) | 0.8173 | 0.64 (0.46-0.89) | 0.0093 |
| Adjusted for HDL | 0.90 (0.52-1.56) | 0.7202 | 0.65 (0.47-0.91) | 0.0125 |
| Adjusted for TG* | 0.80 (0.47-1.36) | 0.4136 | 0.64 (0.46-0.88) | 0.0064 |
| Adjusted for LDL and HDL | 0.81 (0.37-1.77) | 0.5987 | 0.72 (0.43-1.19) | 0.2034 |
| Adjusted for LDL and TG | 1.02 (0.46-2.25) | 0.9557 | 0.91 (0.56-1.46) | 0.6940 |
| Adjusted for HDL and TG | 1.34 (0.73-2.46) | 0.3469 | 0.84 (0.47-1.52) | 0.5728 |
| Adjusted for LDL, HDL and TG | 0.72 (0.41-1.27) | 0.2619 | 0.92 (0.59-1.45) | 0.7304 |
| Adjusted for LDL, HDL, TG and SBP | 0.88 (0.49-1.60) | 0.6847 | 0.94 (0.61-1.44) | 0.7082 |
| Adjusted for LDL, TG and SBP | 0.97 (0.53-1.80) | 0.9346 | 0.88 (0.57-1.36) | 0.5563 |
| Adjusted for LDL, TG, SBP and FSG | 1.63 (0.96-2.77) | 0.0727 | 0.88 (0.58-1.34) | 0.5521 |
| Adjusted for LDL, TG, and FSG | 1.41 (0.85-2.31) | 0.1784 | 0.91 (0.58-1.44) | 0.6992 |

KoGES, Korean Genome Epidemiologic Study; KCPS-II, Korean Cancer Prevention Study-II; BBJ, Biobank of Japan; IVW, inverse variance weighted method; OR, odds ratio; CI, confidence interval; MVMR, multivariable mendelian randomization; LDL, low density lipoprotein; HDL, high density lipoprotein; TG, triglyceride; SBP, systolic blood pressure; FBS, fasting serum glucose

Table 3 showed the crude two sample MR and MVMR results on the effect of indirect bilirubin on ischemic stroke. KoGES results showed a significant negative correlation only in crude two sample MR (P=0.0253). However, in the KCPS-II data analysis, the crude two sample MR showed a borderline significant result, but the MVMR analysis controlling the remaining LDL, HDL, and TG showed a significant result. In addition, MVMR, which simultaneously controlled LDL, HDL, and TG, showed significant results (P=0.0355). This model showed significant results for indirect bilirubin even when SBP or FSG was additionally controlled (right side of Figure 1-C). The F values of indirect bilirubin and control variables used in the model in Table 3 were all above 10 except for SBP, and the assumption as an instrumental variable was satisfied (Supplementary Table 3).

**Table 3.**
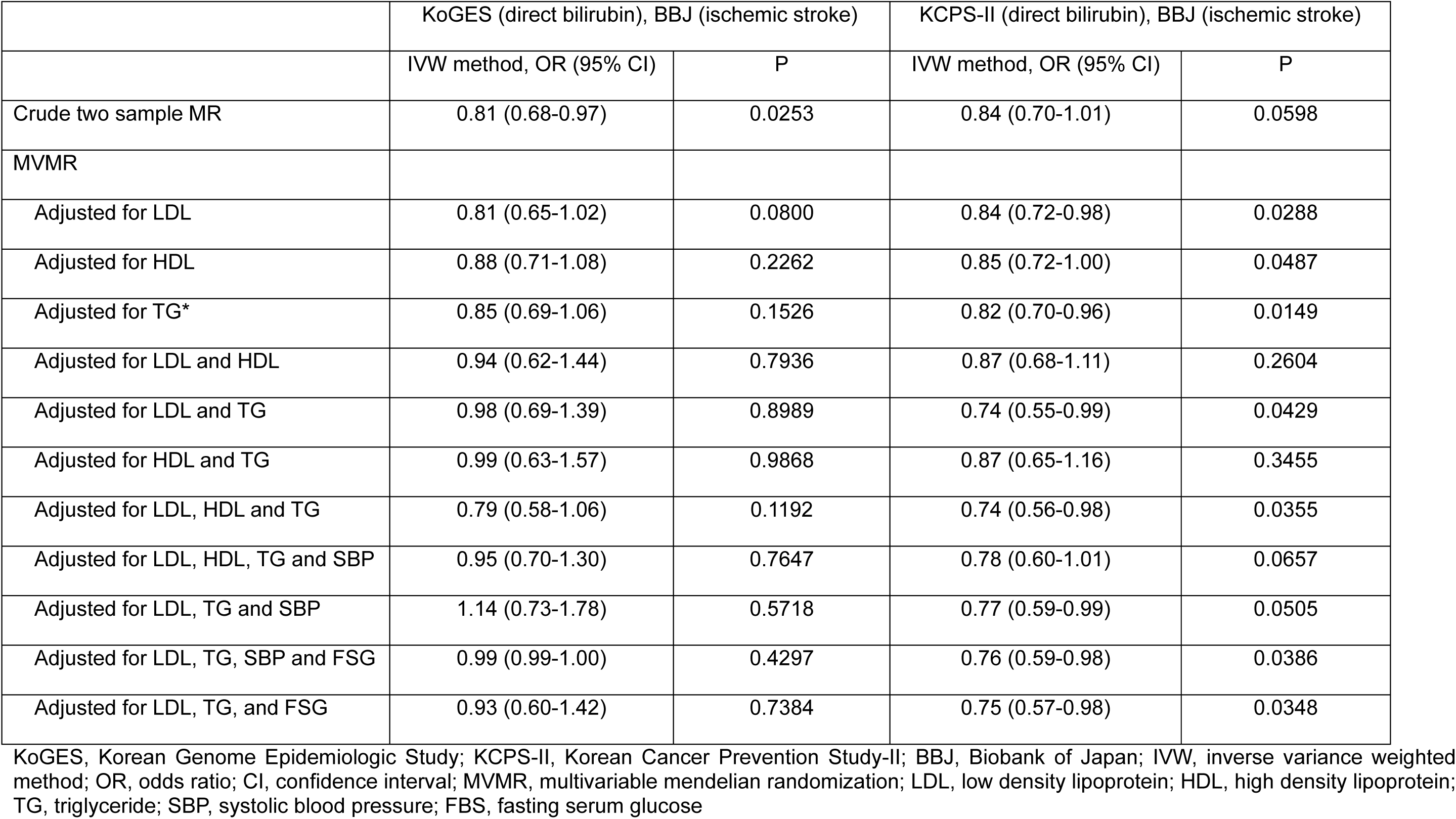
Causal effect of indirect bilirubin on ischemic stroke.

In the models presented in Tables 1, 2, and 3, the common feature of the other control factors except bilirubin was that only SBP consistently showed a significant causal relationship to the risk of ischemic stroke. However, LDL, TG, and FBS were not significant.

## DISCUSSION

In this study, we showed that genetically determined levels of each bilirubin (total, direct, and indirect bilirubin) in serum were causally associated with reduced risk of ischemic stroke in an Asian population. In particular, it was demonstrated that only indirect bilirubin had a strong protective effect in MVMR, which controlled for genetic factors related to ischemic stroke.

This study performed two sample MR to investigate the causal relationship between genetically determined levels of circulating serum bilirubin (total, direct, and indirect bilirubin) and the risk of ischemic stroke. In addition, the causal relationship was shown while controlling for genetic variables of TG, cholesterol (HDL, LDL), SBP, and FBS.

Interestingly, while controlling for genetic variables such as TG, cholesterol (HDL, LDL), SBP, and FBS, indirect bilirubin levels reduced the risk of ischemic stroke. On the other hand, for direct bilirubin, a somewhat weaker association was found. These research results are expected to contribute to understanding the mechanisms caused by oxidative stress in the future.

In the metabolic process of bilirubin, heme is broken down by heme oxygenase to produce carbon monoxide (CO), iron (Fe), and biliverdin.^15,16^ In normal, healthy liver cells, biliverdin is converted to bilirubin via biliverdin reductase A (BVRA). Biliverdin reductase exists as two type of isoenzymes. BVRA and biliverdin reductase B (BLVRB) produce bilirubin IXα and bilirubin IXβ, respectively.^15^ BVRA is critical for adult bilirubin production forming bilirubin IXα, whereas bilirubin IXβ is mostly present during fetal development. Bilirubin IXα is insoluble and binds to albumin in the blood (indirect bilirubin, unconjugated bilirubin), where it is transported to the liver and conjugated.^15^

Afterwards, bilirubin is conjugated in hepatocytes by the UGT1A1 UDP-glucuronosyltransferase enzyme (direct bilirubin, conjugated bilirubin) and is eventually excreted in bile and reaches the intestines.^16,17^

Conjugated bilirubin is isolated by intestinal bacteria and reduced to urobilinoid. In this way, the enzymes and activity sites involved in each bilirubin during the metabolic process are different. Since the manifestation of the disease according to the increase or decrease in each of these levels appears completely different, it is believed that the classification of total bilirubin subgroups will have a significant impact on the results and direction of the research.

In two Sample MR, exposure variables and outcome data are each extracted from two independent biobanks. This is advantageous when exposure and outcome data are not available from the same biobank. This study used serum bilirubin data from KoGES and KCPS-II. For ischemic stroke data, MR analysis could be performed using the same Asian data from BBJ.

Additionally, utilizing multiple samples improves the overall sample size and subsequent precision of causal effect estimates. As a result, we not only re-proved the causal relationship between bilirubin levels and stroke risk reduction in Korean cases in the previous study,^10,11^ but also confirmed the causal relationship between ischemic stroke risk reduction in Japanese as an Asian population.

Evidence from numerous observational studies in humans indicates a strong negative association between serum bilirubin levels and cardiovascular disease. For example, bilirubin has antioxidant properties, including scavenging reactive oxygen species (ROS) and inhibiting nicotinamide adenine dinucleotide phosphate (NADPH) oxidase activity, thereby oxidative stress, which is critically involved in the pathogenesis and development of atherosclerosis.^18,19^

Additionally, serum total bilirubin concentration was negatively associated with arteriosclerosis in Chinese men,^20^ and in a German population, serum bilirubin levels were observed to be inversely associated with coronary artery calcification and cardiovascular disease.^21^In the Chinese population, serum total bilirubin levels were negatively associated with subclinical cerebral infarction, which increased the risk of transient ischemic attack, symptomatic stroke, and cardiovascular disease.^22^ Regarding stroke risk, a cross-sectional study of Americans conducted from 1999 to 2004, the National Health and Nutrition Examination Survey, reported that serum total bilirubin levels were inversely associated with stroke outcomes.^23^ Especially in the Korean population, serum bilirubin concentration was estimated to be negatively correlated with ischemic stroke in men.^6^

Additionally, several experimental studies support the results of this study. When bilirubin acts as a ROS scavenger, indirect bilirubin (albumin-bound unconjugated bilirubin) is oxidized to its non-toxic metabolic precursor biliverdin, which is then recycled back to bilirubin by biliverdin reductase.^19,24,25^ In vitro studies have shown that indirect bilirubin (unconjugated bilirubin bound to albumin) is oxidized by O_2_ −, OH and HO_2_ and that unconjugated bilirubin protects albumin from oxidation by O_2_ −, OH, HO_2_ and ONOO −. Indirect bilirubin can act as an antioxidant by directly scavenging ROS.^19,25,26^ In addition, indirect bilirubin has been shown to inhibit the activity of NADPH oxidase, a major source of ROS in the vascular system, in vitro and in vivo, suggesting that the production of ROS is reduced by indirect bilirubin.^18,27^ Moreover, bilirubin has been shown to interact with other antioxidants to synergistically inhibit lipid peroxidation.^28,29^ Taken together, these experimental findings indicate that indirect bilirubin can function as an antioxidant by interacting synergistically with other antioxidants to scavenge ROS, inhibit NADPH oxidase activity, and inhibit lipid oxidation.

Bilirubin is a key indicator that continues to be studied today as studies have linked it to the risk of oxidative stress-related diseases in humans, such as cardiovascular disease, including stroke, diabetes, metabolic syndrome, certain cancers, and autoimmune diseases.^29^ The above research results suggest that specific studies of other oxidative stress-related diseases can be conducted based on the detailed bilirubin index and each bilirubin level presented by controlling various genetic variables.

The strengths and limitations of this study are as follows. The strength is that bilirubin data was obtained from KCPS-II and KoGES data as a Korean biobank for two-sample MR analysis, and stroke data is large-scale data using the Japanese biobank BBJ. Additionally, unlike previous studies, this is the first study to look at the effects of direct and indirect bilirubin in addition to total bilirubin. A limitation is that only ischemic stroke was included in the study, and total stroke and hemorrhagic stroke were not included. However, in previous studies examining bilirubin and stroke, only ischemic stroke showed a significant association.

## CONCLUSIONS

This study causally proved that indirect bilirubin is a very significant causal protective factor in the risk of ischemic stroke. However, when comparing systolic blood pressure and indirect bilirubin, which are shown to be the same causal factor although the direction is different, indirect bilirubin has a very wide confidence interval. This implies that the reliability and statistical significance of the estimation results remain low. In the future, it is believed that research should continue to gain a deeper understanding of the protective effect of indirect bilirubin on ischemic stroke.

## Data Availability

Data availability statement When a paper is published, summary data are available.

## Acknowledgements

We would like to thank the staff at Korea Medical Institute for their assistance in data collection. We would also like to thank the staff at the other 11 general health examination centers who participated in this study for their assistance in data collection.

## Contributors

SHJ, KJJ and HK had full access to all of the data in the study and takes responsibility for the integrity of the data and the accuracy of the data analysis. Study concept and design: SHJ, KJJ and HK. Diagnostic testing and statistical analysis: SHJ and KJJ.

Drafting of the manuscript: JWS and KJJ.

Critical revision of the manuscript for important intellectual content: SHJ and HK. Study supervision and organization of the project: SHJ and HK.

## Ethical approval

This study protocol was approved by the Institutional Review Board of the Severance Hospital (approval number: 4-2011-0277).

